# Whole genome sequence analysis of *Salmonella* Typhi provides evidence of phylogenetic linkage between cases of typhoid fever in Santiago, Chile in the 1980s and 2010-2016

**DOI:** 10.1101/2022.01.19.22269577

**Authors:** Mailis Maes, Michael J. Sikorski, Megan E. Carey, Ellen E. Higginson, Zoe A. Dyson, Alda Fernandez, Pamela Araya, Sharon M. Tennant, Stephen Baker, Rosanna Lagos, Juan Carlos Hormazábal, Myron M. Levine, Gordon Dougan

**Author notes:** These authors contributed equally to this work (Mailis Maes, Michael J. Sikorski). These authors contributed equally to this work (Myron M. Levine, Gordon Dougan).

## Abstract

Typhoid fever epidemiology was investigated rigorously in Santiago, Chile during the 1980s, when *Salmonella enterica* serovar Typhi (*S*. Typhi) caused seasonal, hyperendemic disease. Targeted interventions reduced the annual typhoid incidence rates from 128-220 cases/10^5^ population occurring between 1977-1984 to <8 cases/10^5^ from 1992 onwards. As such, Santiago represents a contemporary example of the epidemiologic transition of an industrialized city from amplified hyperendemic typhoid fever to a period when typhoid is no longer endemic. We used whole genome sequencing (WGS) and phylogenetic analysis to compare the genotypes of *S*. Typhi cultured from acute cases of typhoid fever occurring in Santiago during the hyperendemic period of the 1980s (n=74) versus the nonendemic 2010s (n=80) when typhoid fever was rare. The genotype distribution between “historical” (1980s) isolates and “modern” (2011-2016) isolates was similar, with genotypes 3.5 and 2 comprising the majority of isolations, and 73/80 (91.3%) of modern isolates matching a genotype detected in the 1980s. Additionally, phylogenomically ‘ancient’ genotypes 1.1 and 1.2.1, uncommon in the global collections, were also detected in both eras, with a notable rise amongst the modern isolates. Thus, genotypes of *S*. Typhi causing acute illness in the modern nonendemic era match the genotypes circulating during the hyperendemic 1980s. The persistence of historical genotypes may be explained by chronic typhoid carriers originally infected during or before the 1980s.

**Author summary:** Studies of *Salmonella* Typhi (the cause of typhoid fever) rarely include isolates collected both before and after the interruption of hyperendemic transmission because this typically occurred decades before modern bacteria preservation methods. After interruption, it was assumed that sporadic cases and infrequent outbreaks were due to either chronic biliary carriers or importations, but this was difficult to characterize with low resolution bacterial typing methods. In Santiago, Chile, typhoid fever persisted at hyperendemic levels through the 1980s until organized control efforts in the 1980s and changes to wastewater policy in 1991 caused annual typhoid incidence to plummet.

In this study, we used whole genome sequencing (WGS) to investigate whether recent sporadic cases occurring in Santiago in the 2010s were genomically similar to *S*. Typhi circulating in the 1980s, or dissimilar, possibly representing importations of *S*. Typhi from outside of Chile. We found concordance amongst *S*. Typhi genotypes between the 1980s and 2010s, and differences from genotypes circulating in Southeast Asia and Africa where typhoid remains hyperendemic. Our findings suggest that a proportion of modern, rare typhoid cases in Santiago are autochthonous, and that chronic carriers or another unknown reservoir likely contribute. Broadly, our findings corroborate the epidemiologic importance of long-term reservoirs of typhoid fever decades after typhoid elimination.

## Introduction

Typhoid fever is a systemic infection, caused by the bacterium *Salmonella enterica* subspecies *enterica* serovar Typhi (*S*. Typhi), which is spread through ingestion of food and water contaminated with human faeces containing *S*. Typhi. Typhoid fever in Santiago, Chile rose to hyper-endemic levels in the mid-1970s with a striking annual incidence of 177-220 cases per 10^5^ population per year from 1977-1983, partially declined during the years 1984-1991 (51-128 cases/10^5^ per year), and finally plummeted below 8 cases/10^5^ per year after 1991, reaching nonendemic levels of ∼0.5/10^5^ in 2012 [1]. During the hyperendemic years, the highest incidence rate was observed in school age children, and case numbers peaked in the rainless summer months [2]. To address this public health issue, in 1979 the Ministry of Health of Chile implemented a Typhoid Fever Control Program involving strategic preventive interventions including large-scale field trials of live oral typhoid vaccine Ty21a in >500,000 schoolchildren [3-6], and identification and treatment of chronic carriers among food handlers [7]. Collectively, these interventions led to a steady decline in typhoid cases between 1983 and 1990 [3-6, 8].

Epidemiologic and environmental bacteriology studies in the 1980s undertaken by the Chilean Typhoid Fever Control Program also incriminated the use of raw untreated sewage water for irrigation of vegetable crops during the rainless summer months as a key risk factor responsible for amplified transmission of *S*. Typhi in Santiago [8, 9]. In April 1991, the government of Chile abruptly prohibited the practice of irrigating crops with untreated sewage wastewater in Santiago [1, 8, 10, 11]. This action was taken following the enormous explosive epidemics of El Tor cholera in 1991 that occurred in neighbouring countries to the North, including in Peru, beginning in late January [12, 13], followed by Ecuador in February [13], and Colombia in March [13]. The pivotal stimulus for the intervention was a small outbreak of cholera in Santiago, Chile, in April, 1991 [1, 8, 10, 11]. Following imposition and strict enforcement of the prohibition of use of untreated sewage water for irrigation, the Santiago cholera epidemic quickly came to an end [10, 11], and the incidence of typhoid dropped precipitously in subsequent years [1, 8, 10]. From 2000-2014, the annual incidence of confirmed typhoid fever in Santiago has ranged from 0.20-1.18 cases/10^5^ population. The current rare sporadic cases of typhoid in Santiago include illnesses among: travellers who visit countries with high incidence rates of typhoid; immigrants from typhoid-endemic countries; and transmission from Chilean chronic carriers who were originally infected decades earlier (pre-1991) during the era of hyperendemic transmission.

Phylogenetic analysis of whole genome sequencing (WGS) data has become the gold standard for inferring relatedness between *S*. Typhi isolates [14]. WGS can also be used to assign genotypes, identify and characterize plasmids, compare gene content and homology, and identify molecular determinants of antibiotic resistance and virulence [15, 16]. Whereas WGS data from many global isolates are available, few bacterial collections include isolates from both before and after the interruption of amplified hyperendemic typhoid fever. Also, WGS-derived information concerning the genotypes isolated from typhoid fever cases in South American countries is scarce. The majority of publicly available sequenced isolates come from hyperendemic regions in South Asia and East Africa where H58 (4.3.1) genotypes dominate [17].

Herein we describe a genomic analysis of *S*. Typhi isolates from Santiago, Chile from two distinct epidemiologic periods including the early 1980s (N=74), when typhoid was hyperendemic, and from 2010-2016 (N=80), a quarter century after the interruption of amplified transmission of *S*. Typhi in Santiago. We show that the genotypic composition of the population structure is similar between these two periods, even though the local epidemiology has changed drastically. In addition, we describe a potential carrier-related cluster of the “ancient” 1.1 clade, and the identification of a new variant of plasmid pHCM2 carried by 2 isolates of another ancient genotype (1.2.1).

## Material and methods

### Study design

In total, 74 “historical” isolates of *S*. Typhi from cases of acute typhoid fever in Santiago, Chile in the period 1981 to 1986, during the era of hyperendemic typhoid, were obtained from the culture collection of the Center for Vaccine Development and Global Health of the University of Maryland, Baltimore. “Modern” *S*. Typhi isolates (n=80) were strains collected from acute cases of typhoid fever and referred to the Instituto de Salud Pública de Chile (ISP, Institute of Public Health) by clinical microbiology laboratories from 2011-2016; these referred strains were generally accompanied by only limited demographic and clinical information.

### DNA extraction and sequencing

DNA was isolated from the 154 *S*. Typhi isolates using the Norgen DNA extraction kit as per the manufacturer’s instructions. Genomic DNA was sequenced by Illumina HiSeq at the Wellcome Sanger Institute, generating 125 bp paired reads. Raw read data were deposited in the European Nucleotide Archive and individual accession numbers are listed in Table S1.

### Assembly, mapping, SNP-calling and genotype assignment

Raw Illumina reads were assembled using Velvet v1.2 via the Wellcome Sanger Institute automated analysis pipeline [18]. Sequenced reads were mapped and annotated, single nucleotide polymorphisms (SNPs) were called against the *S*. Typhi CT18 reference genome [19] using SMALT v0.7.4 and subsequent variant calling was carried out as previously described [20]. Genotypes were assigned using GenoTyphi (v1.9.0) [14, 16].

### Phylogenetic analysis

Previously defined [21-23] recombinant regions such as prophages and plasmids were excluded manually, and any remaining recombinant regions were filtered using Gubbins (v2.5.0) [24]. The resultant core genome alignment was used to infer Maximum Likelihood (ML) phylogenies using RAxML (v8.2.8) [25], specifying a generalized time-reversible model and a gamma distribution to model site-specific rate variation (GTR+ Γ substitution model; GTRGAMMA in RAxML) with 100 bootstrap pseudoreplicates used to assess branch support. SNP distances for the core genome alignment of all the strains have also been calculated from this alignment using snp-dists package (available at: https://github.com/tseemann/snp-dists). Resulting phylogenies were visualized with iTOL [26] and are available for interactive inspection on Microreact [27] (https://microreact.org/project/j25cGgoVX8ixQCRstPVhgo-chile-1980-2010).

### Molecular determination of AMR, plasmids and virulence genes

Raw read data were screened using SRST2 v0.2.0 [15] with the ARG-ANNOT [28], PlasmidFinder [29], and *Salmonella* Virulence Factor DB databases [30] to detect known and putative antimicrobial resistance (AMR) genes, known plasmid replicons, and known virulence-associated genes, respectively. Homology cutoffs of 90% nucleotide similarity were used for all SRST2 screens.

Plasmid pHCM2 sequences were assembled with Unicycler [31], and the circularised sequences extracted from *de bruijn* assembly graphs with Bandage (v0.8.1) [32]. These were then compared with the pHCM2 reference sequence (accession no: AL513384) using Artemis-ACT [33] and visualized with EasyFig [34].

## Results

### *S*. Typhi genotypes in Santiago, Chile

Genotypic analysis indicated that most of the isolates from Santiago in this study belonged to genotypes 2 (n=43, 27.9%), 3.5 (n=51, 33.1%) and 1.1 (n=22, 14.3%) (Table 1). While these three genotypes were the most prevalent, we also detected 11 additional distinct genotypes across the two time periods. These include three previously described importations of isolates belonging to H58 genotypes 4.3.1.1 and 4.3.1.2 [35].

**Table 1.** S. Typhi genotypes detected in isolates from acute clinical cases in Santiago, Chile during the hyperendemic 1980s versus the nonendemic 2010s.

|  | 1983-1986 | 2011-2016 | TOTAL |
| --- | --- | --- | --- |
| **1.1** | 2 (2.7%) | 20 (25%) | 22 (14.3%) |
| **1.2.1** | 1 (1.4%) | 5 (6.3%) | 6 (3.9%) |
| **2** | 30 (40.5%) | 13 (16.3%) | 43 (27.9%) |
| **2.0.2** | 11 (14.9%) | 5 (6.3%) | 16 (10.4%) |
| **2.2** | - | 3 (3.8%) | 3 (1.9%) |
| **2.3.2** | 1 (1.4%) | 1 (1.3%) | 2 (1.3%) |
| **2.3.3** | 1 (1.4%) | 1 (1.3%) | 2 (1.3%) |
| **2.3.4** | 1 (1.4%) | - | 1 (0.6%) |
| **3.1** | 1 (1.4%) | - | 1 (0.6%) |
| **3.3** | - | 1 (1.3%) | 1 (0.6%) |
| **3.5** | 25 (33.8%) | 26 (32.5%) | 51 (33.1%) |
| **4.1** | 1 (1.4%) | 2 (2.5%) | 3 (1.9%) |
| **4.3.1.1** | - | 2 (2.5%) | 2 (1.3%) |
| **4.3.1.2** | - | 1 (1.3%) | 1 (0.6%) |
| **TOTAL** | 74 | 80 | 154 |

The genomic population structure of *S*. Typhi in Santiago appeared highly consistent between the hyperendemic and post-hyperendemic eras (Fig 1). Both rare “ancient” genotypes 1.1 and 1.2.1, as well as more commonly found genotypes 2 and 3.5, were frequently present across both study periods. Despite several decades having passed since the cessation of the era of hyperendemic transmission, most of the post-2010 sporadic *S*. Typhi isolates shared bootstrap-supported phylogenetic clusters with the 1980 hyperendemic isolates. Only within genotype 1.1 did recent and historical sequences form independent clusters. The intermingling of historical and recent sequences suggests that the 2010s isolates originated from progenitors within the same pool of *S*. Typhi circulating in the 1980s.

**Fig. 1.**
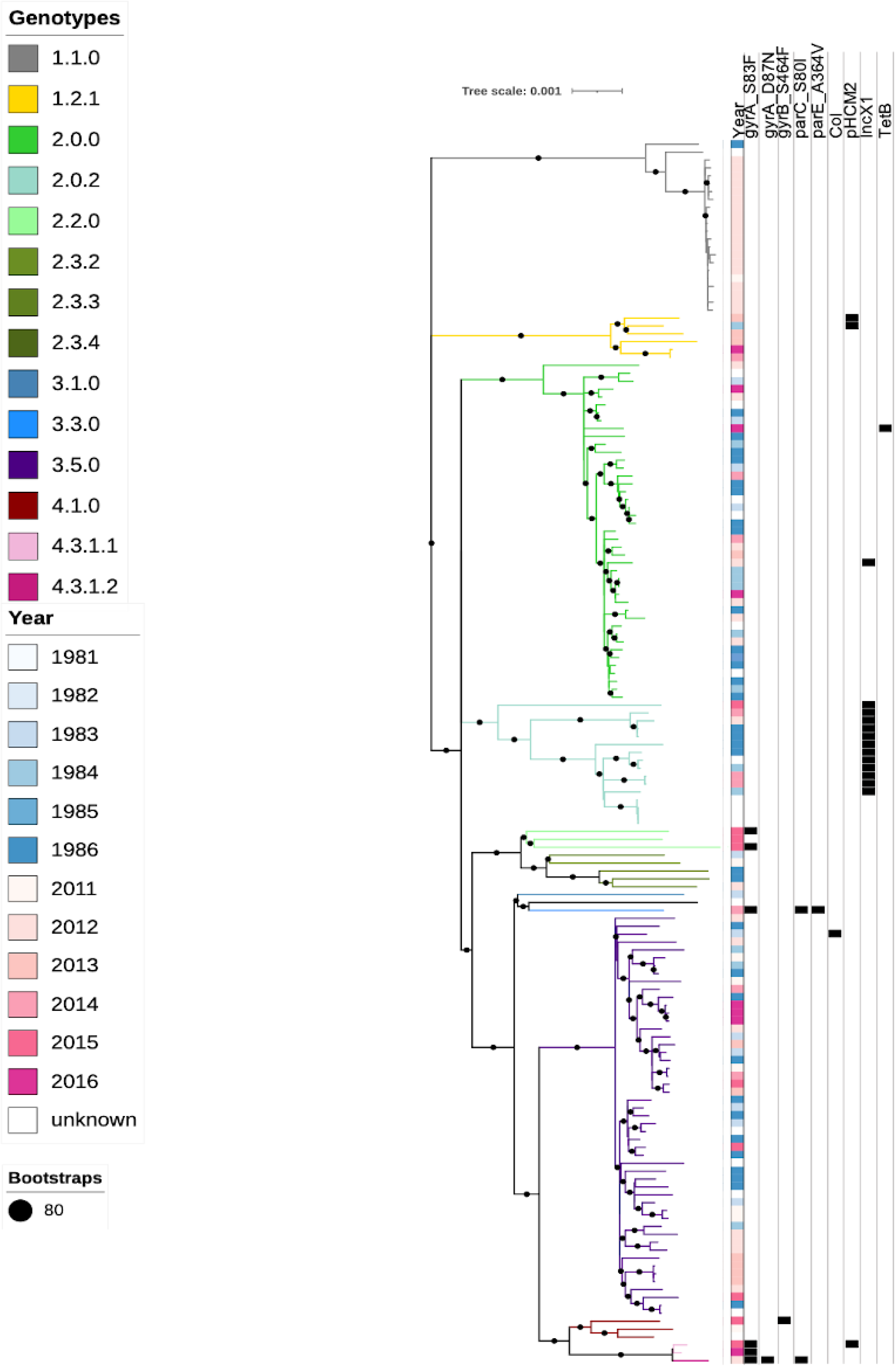
Genotypes and molecular determinates of AMR and plasmids in the Santiago collection. SNP based maximum likelihood core genome phylogeny indicating the genotypes of all the Santiago *S*. Typhi isolates is shown by the branch colour as per the inset legend. A diverse range of 14 genotypes were detected including 1.1, 1.2.1, 2, 2.0.2, 2.2, 2.3.2, 2.3.3, 2.3.4, 3.1, 3.3, 3.5, 4.1, 4.3.1.1, 4.3.1.2. Most of the isolates belonged to genotype 1.1, 2 and 3.5. Year of isolation, SNPs in the QRDR region associated with reduced susceptibility to fluoroquinolones as well as plasmids and detected resistance genes are shown in the columns adjacent to the tree. Bootstrap confidence of 80 or above is indicated with a black circle on the branch.

Amongst the 151 non-H58 isolates, few antimicrobial resistance mechanisms were detected. One genotype 2.0.0 isolate carried a *tetB* gene, known to encode a mechanism of resistance to tetracyclines (albeit not an antibiotic of clinical relevance for treatment of typhoid fever). Two genotype 2.2 isolates and one genotype 4.1 isolate had single non-synonymous point mutations in the Quinolone Resistance Determining Region (QRDR) of genes *gyrA* and *gyrB*, associated with reduced susceptibility to fluoroquinolones [36]; in the late 1980s, ciprofloxacin was the antibiotic of choice for treating patients with typhoid fever. Only 17 candidate plasmids were detected across all the isolates, including a Colicin plasmid, three related to plasmid pHCM2 and 13 of incompatibility type IncX1 (Fig 1).

### Global transmission dynamics

To better understand how this local population structure fits into the global context of *S*. Typhi, a maximum likelihood phylogenetic tree was generated using a total of 2,013 non-H58 *S*. Typhi genomes from approximately 70 countries (Fig 2). The *S*. Typhi isolates from Santiago of genotypes 3.5, 2, 1.1 and 1.2.1 clustered together and were genetically distinct from those sequenced previously from other countries. The other genotypes present in Santiago were mostly found mostly on long branches, indicating significant differentiation from their closest relatives in the global context phylogeny.

**Fig. 2.**
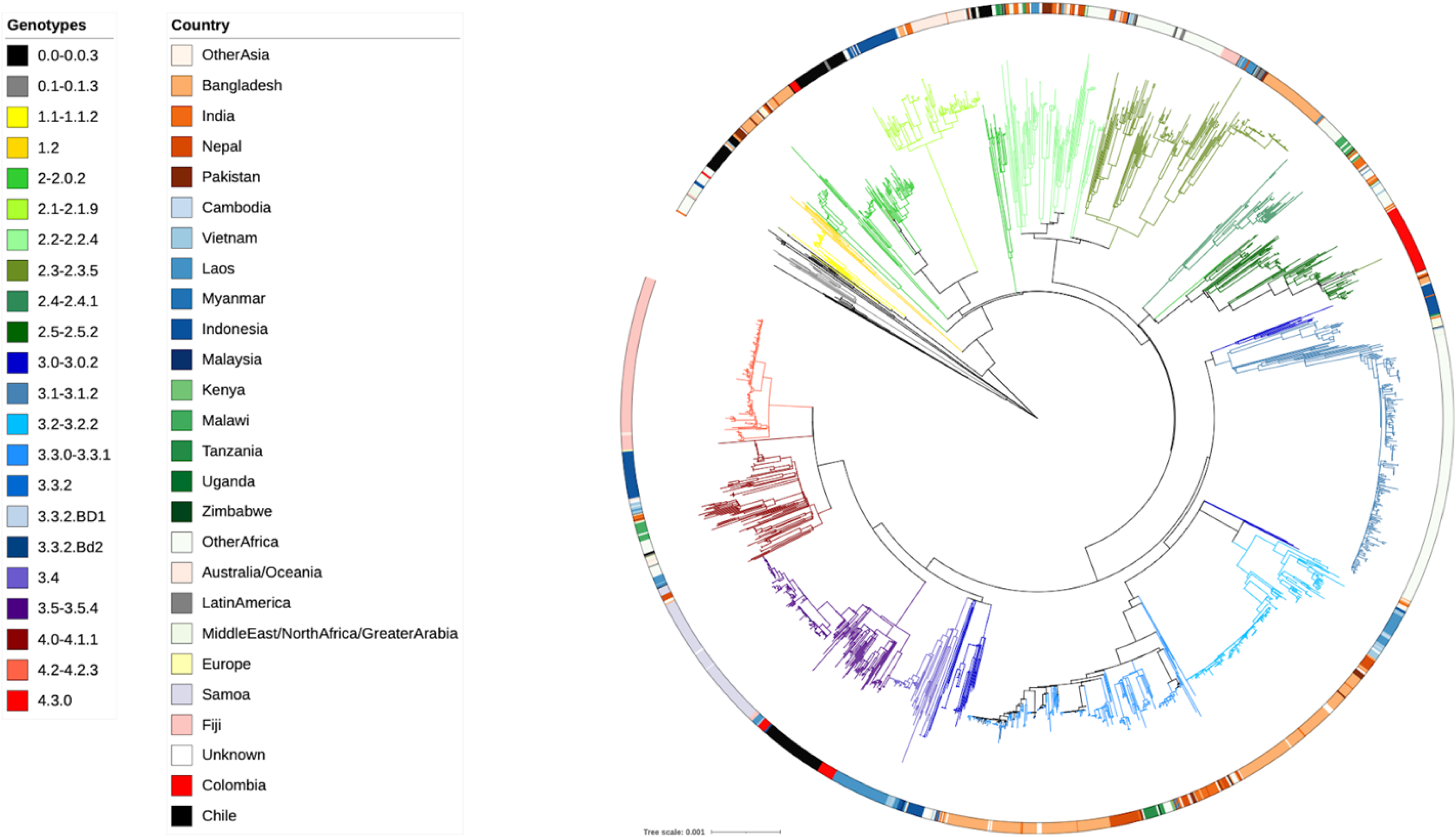
A global context maximum likelihood core genome phylogeny incorporating publicly available non-H58 and Santiago *S*. Typhi isolates. SNP based core genome maximum likelihood tree of 2013 global isolates, branches are coloured by genotype as per the inset legend; the ring indicates country of isolation as per the inset legend; Santiago isolates are highlighted in black and the Colombian isolates in red.

### “Ancient” *S*. Typhi clades endemic to Chile

Two of the clades common in our dataset, 1.1 and 1.2, are found closer to the root of the global *S*. Typhi tree and thus can be considered to comprise “ancient” clades, which are detected with very low global frequency. Of the 1.1 isolates (n=22), two were from the 1980s collection (Fig 3). Only six other isolates, 0.3% of the global collection described by Wong et al [14], belonged to clade 1.1. Interestingly, a cluster of closely related genotype 1.1 *S*. Typhi isolates was detected in the years 2011 (1 isolate) and 2012 (19 isolates). These isolates also exhibit lower genetic variation (median SNP distance of 8) than those observed among the two 1980s 1.1 isolates (119 SNPs). The smallest SNP distance between a 1980s 1.1 isolate and a 2010s 1.1 isolate was 79 SNPs. This suggests the 2011-2012 isolates have a recent common ancestor.

**Fig. 3.**
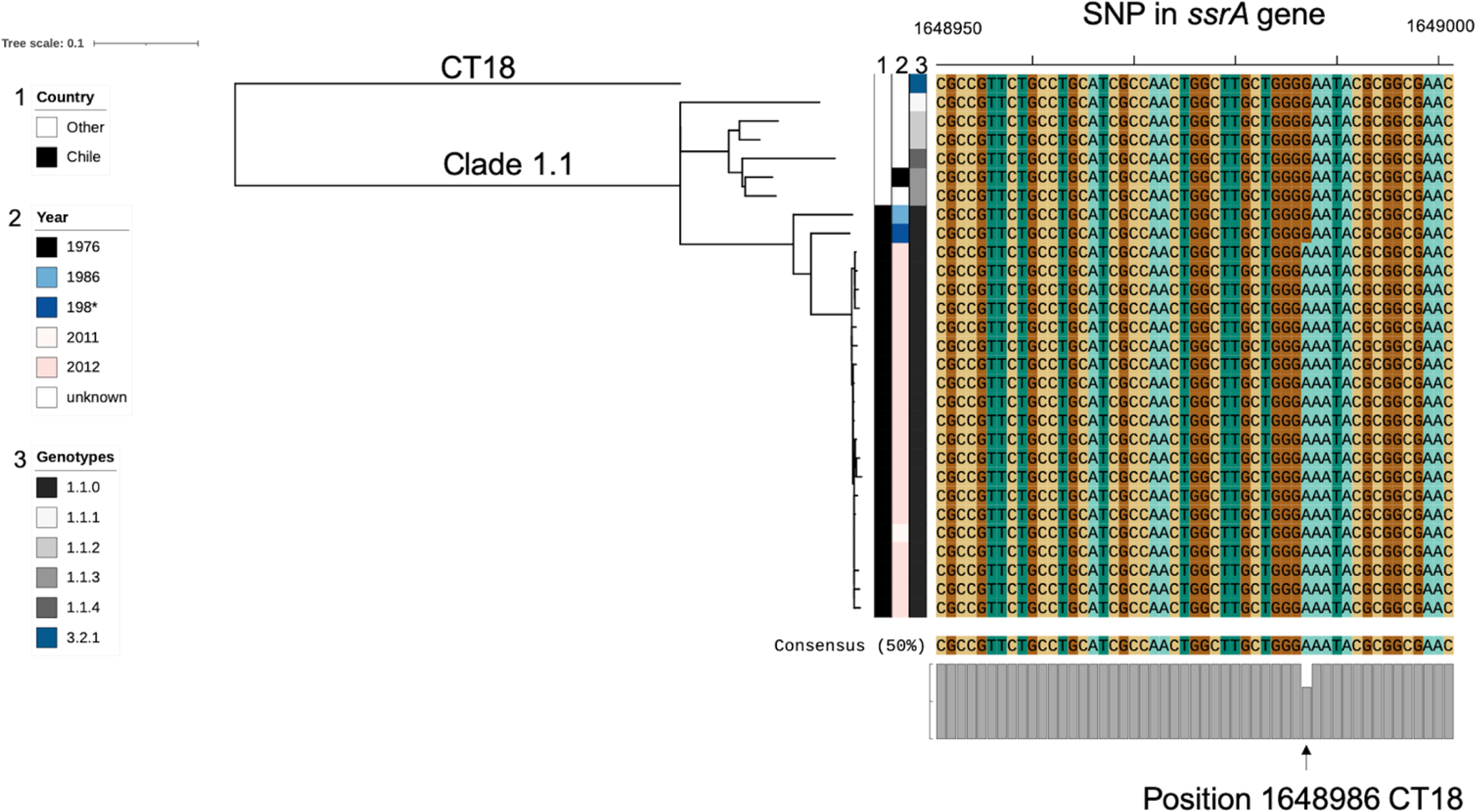
*S*. Typhi clade 1.1 phylogeny showing sequences carrying *ssrA* mutation (*ssrA-* G639E). Phylogenetic core genome analysis of all *S*. Typhi isolates of genotype 1.1 identified from the global collection. The majority of these isolates were collected in Santiago (black bar), two isolates belonged to the 1980s collection appearing ancestral to the 2010s collection. The twenty isolates from 2011-2012 showed an interesting cluster with a median SNP distance of 8 SNPs which is unlikely to have evolved among acute cases, where the mutation rate is calculated to be < 1 SNP/genome/year, in one year. Also shown is an alignment of a section of the *ssrA* gene, showing the conserved SNP at position 1648986 of the *S*. Typhi CT18 reference genome. * the precise isolation year of one of the 1980s isolates was unclear.

We sought to determine if there was a specific virulence gene associated with the 2011-2012 genotype 1.1 cluster, using SRST2 with the *Salmonella* VFDB. Although there were no additional or missing virulence factor-associated genes detected, this screening did reveal that all of the genotype 1.1 isolates from 2011-2012 cluster harboured a SNP in the *ssrA* gene. The alignment of this gene revealed the same missense mutation (G to A at position 1648968 of CT18, causing a glycine to glutamic acid amino acid change) in the *ssrA* gene (*ssrA-* G639E), (Fig 3).

The other ancient clade present amongst the Santiago isolates was genotype 1.2.1. This was the only clade in our datasets (except from one recently imported H58 isolate) in which isolates harboured pHCM2 plasmids. pHCM2 plasmids are found in ∼15% of *S*. Typhi and are generally highly conserved (Fig 4A). To our knowledge based on the data available in the *S*. Typhi public collection on Pathogenwatch: (https://pathogen.watch/collection/1y3cdsjq55hf-public-genomes-accessedDec2021) pHCM2 plasmids have not been detected in ancient clades 0.0 to 1.2 previously. Two pHCM2 plasmids were found among our isolates from genotype 1.2.1; one in a 1984 isolate (ERR2710544) and the other from 2013 (ERR4289185). These plasmids were highly similar to each other, but significantly different to the pHCM2 found on *S*. Typhi reference CT18 (accession no: AL513384), and to another novel pHCM2 found in a 1973 isolate from Thailand [37]. The pHCM2 sequences described here differed by >2000 SNPs (98.4% nucleotide identity, 89% coverage) from the reference (Fig 4).

**Fig. 4.**
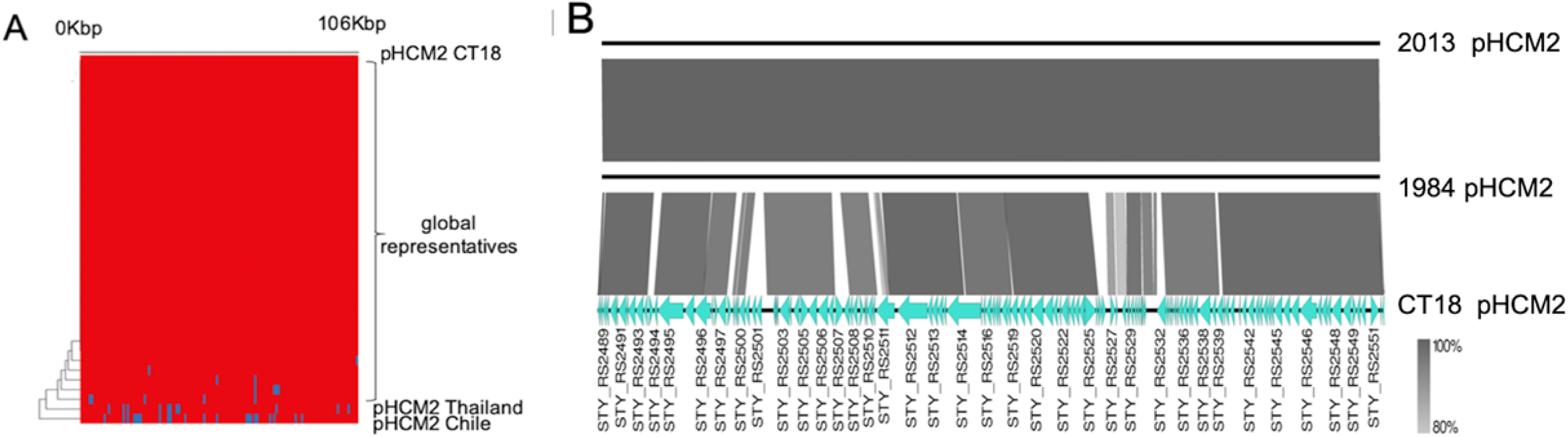
Genetic comparison of cryptic plasmid pHCM2-like sequences. Global representative collection of pHCM2 aligned to the reference pHCM2 from CT18. (A) Presence and absence of genes located on pHCM2. Red indicates presence of the genes located on the reference pHCM2 sequence from CT18, while blue indicates absence of the genes compared to the reference. Nucleotide BLAST comparison of the 2 Chilean assembled pHCM2-like plasmids (ERR271054, ERR4289185) against the reference pHCM2 plasmid from *S*. Typhi CT18 (accession no:AL513384) (B). Gene annotations are shown for the reference pHCM2 from CT18, with the shading intensity indicating percent nucleotide homology between the plasmid sequences.

## Discussion

This study utilizes WGS analyses to compare *S*. Typhi isolates from patients who developed acute typhoid fever in Santiago during the modern era (i.e., 2011-2016, decades post-interruption of amplified typhoid transmission in 1991) with isolates from acute cases during the hyper-endemic era of the 1980s. Using phylogenetics and genome-wide screening, we compared the genotypes, AMR determinants, plasmid content of *S*. Typhi isolates between these two time periods. Whilst comparisons of isolates from the 1980’s with isolates from 1990 (post vaccinating interventions-but pre-irrigation ban, when incidences were still high 47 cases /10^5^ [1]) have been undertaken previously with pulse field gel electrophoresis (PFGE) [38], to our knowledge, this is the first WGS-based analysis comparing *S*. Typhi collected during a period of hyper-endemicity and decades after typhoid control.

The observation that the 1980s and 2010s collections of *S*. Typhi isolates do not cluster independently within our commonly found genotypes (2, 3.5 and 1.2.1) suggests that many of the sporadic cases detected in the 2010s share common ancestry with *S*. Typhi cases from the 1980s. These may, in turn, be linked to shedding of *S*. Typhi from chronic typhoid carriers. This is expected decades after hyper-endemic typhoid is controlled, as chronic carriers will continue to shed sporadically until they are treated or die out of the population [39].

Although there have been several detected introductions of the globally dominant H58 *S*. Typhi clade (4.3.1) into Santiago, as occurs with most industrialized countries in North America, Europe, and Australia, this was not a frequent genotype found in the sporadic modern Chilean cases. This is distinct from countries such as the United Kingdom where imported cases of H58 *S*. Typhi have become common among travel-associated typhoid cases in recent years [40, 41].

*S*. Typhi WGS data from a recent study in Colombia found genotypes 2, 2.5 and 3.5 to be the local endemic genotypes [42]. Genotype 2.5 was not detected among the Santiago isolates; however, genotype 3.5 and 2 were detected in Santiago, and were the nearest genomic neighbours of the Colombian isolates on the global tree. Even though the sequenced isolates from Colombia and Santiago, Chile both contained isolates form the 2010s and historical isolates, they clustered separately on the global tree, suggesting that there has not been obvious transmission between the two countries. More representative WGS data from other Central and South American countries are needed to determine if this is the case with other unsampled endemic countries in the area.

One of the more interesting findings from this dataset was the cluster of 20 highly related strains of genotype 1.1 in 2011-2012. These isolates differed by a median of 8 SNPs in the core genome. For context, the average core genome mutation rate in *S*. Typhi is approximately 1 substitution/genome/year every year [21, 43-45]. As such, it is unlikely that these isolates originated from a clonal source and sequentially acquired this genetic variation in <2 years. Conversely, these strains are clearly highly related and were all collected during a short time span. A potential explanation to account for this cluster is that it was an outbreak caused by bacteria shed from a long-term carrier. *S*. Typhi colonizing the gallbladder or other long-term physiologic niche of a chronic typhoid carrier may evolve and mutate over time into a heterogeneous, yet closely related, community of bacteria [46]. Thus, the closely related nature of these isolates occurring mainly over a single year and during a period when typhoid was no longer hyperendemic in Santiago could be explained by an outbreak instigated by a chronic carrier who was shedding multiple highly related strains. If this hypothetical carrier was working as a food handler, intermittently contaminating food vehicles consumed by subjects during 2011-12, this could result in the cluster of closely related isolates we observe. The cessation of cases could then be explained by a change of employment or residence, a cholecystectomy procedure, a course of antibiotics that concentrate in the gallbladder (e.g., ciprofloxacin) prescribed for another infection or surgery, or death of the hypothetical carrier at the end of 2012.

Notably, the only detectable difference in virulence factor-associated genes from this potential carrier-related 2011-2012 cluster is the *ssrA* missense mutation. The *ssrA/B* two component response regulator is a principle regulator of *Salmonella* pathogenicity island-2 (SPI-2) [47, 48]. SPI-2 has been shown to be insignificant in *S*. Typhi macrophage survival capability and *S*. Typhi’s infection efficiency of humanized mice [49, 50]. The *ssrA* gene has been linked to biofilm formation in *Salmonella* and persistence on gallstones [51, 52]. This observation may be worth additional investigation to investigate the role of SPI-2 and *ssrA* in Typhoid carriage.

Our analysis identified a novel variant of cryptic plasmid pHCM2 in two sequences from genotype 1.2.1. A pHCM2 type plasmid was present in the *S*. Typhi reference isolate CT18 from Vietnam [53] and the plasmid family was described in some detail [54]. Interestingly, any role for pHCM2 in *S*. Typhi biology remains elusive. We know that almost all of the pHCM2 plasmids in the global *S*. Typhi collection are highly conserved in terms of overall nucleotide diversity. However, the Chilean pHCM2 from clade 1.2.1 is substantially different from most other family members. The only other reported pHCM2 plasmid with a similar level of variation is harboured in a *S*. Typhi Thailand isolate from 1973 [55].

In conclusion, this genomics analysis suggests that asymptomatic typhoid carriers (or other unknown reservoirs) are likely responsible for continuing sporadic typhoid cases in Chile. The ancient clades present in the country have persisted. We recommend that all sporadic typhoid cases and clusters should be evaluated using epidemiological outbreak investigations and WGS as routine surveillance. This will help ensure the detection of any potential introduction or development of drug-resistant *S*. Typhi and may lead to the identification and treatment of carriers, which would further limit ongoing disease transmission.

## Data Availability

All Raw Sequencing files are available from the ENA database (accession numbers are listed in Table S1).

## Acknowledgements

We express our thanks to the typhoid fever patients whose samples were analysed and included in this paper, as well as the local public health personnel in Chile. Furthermore, we wish to thank Nicholas Thomson for access to the Sanger analysis pipelines.

## Authorship declaration

All the Co-authors contributed to manuscript editing

## Authors contributions

**Conception and design of experiments**

Myron M. Levine, Rosanna Lagos,

**Sample collection and DNA extractions**

Alda Fernandez, Pamela Araya, Rosanna Lagos, Juan Carlos Hormazabal

**Analyzed the data**

Mailis Maes, Megan E. Carey, Ellen E. Higginson,

**Contributed reagents/materials/analysis tools**

Zoe A. Dyson, Stephen Baker, Michael J. Sikorski, Sharon M. Tennant,

**Funding acquisition**,

Myron M. Levine, Gordon Dougan

**Writing original draft**

Mailis Maes, Michael J. Sikorski, Megan E. Carey,, Ellen E. Higginson

**Writing review and editing**

Mailis Maes, Michael J. Sikorski, Megan E. Carey, Ellen E. Higginson,, Myron M. Levine, Zoe A. Dyson, Sharon M. Tennant

## Data availability

Raw sequence data have been submitted to the European Nucleotide Archive (ENA) and accession numbers of individual isolates are listed in the appendix.

